# Clinical features of COVID-19 patients in Abdul Wahab Sjahranie Hospital, Samarinda, Indonesia

**DOI:** 10.1101/2020.05.27.20114348

**Authors:** Swandari Paramita, Ronny Isnuwardana, Marwan Marwan, Donny Irfandi Alfian, David Hariadi Masjhoer

**Affiliations:** Center of Excellence for Tropical Studies, Mulawarman University, Samarinda, Indonesia; Faculty of Medicine, Mulawarman University, Samarinda, Indonesia; Abdul Wahab Sjahranie Hospital, Samarinda, Indonesia

## Abstract

**Introduction:** Coronavirus Disease (COVID-19) is caused by SARS-CoV-2 infection. Indonesia officially established the first COVID-19 confirmation case in early March 2020. East Kalimantan has been determined as a candidate for the new capital of Indonesia since 2019. This makes Abdul Wahab Sjahranie Hospital Samarinda as the largest hospital there has been designated as the main referral hospital for COVID-19 patients in East Kalimantan. We report the epidemiological, clinical, laboratory, and radiological characteristics of these patients.

**Methods:** All patients with laboratory-confirmed COVID-19 by RT-PCR were admitted to Abdul Wahab Sjahranie Hospital in Samarinda. We retrospectively collected and analyzed data on patients with standardized data collection from medical records.

**Results:** By May 8, 2020, 18 admitted hospital patients had been identified as having laboratory-confirmed COVID-19. Most of the infected patients were men (16 [88.9%] patients); less than half had underlying diseases (7 [38.9%] patients). Common symptoms at the onset of illness were cough (16 [88.9%] patients), sore throat (8 [44.4%] patients), and fever (8 [44.4%] patients). Laboratory findings of some patients on admission showed anemia. Levels of aspartate aminotransferase (AST) and alanine aminotransferase (ALT) were increased in 10 (55.6%) of 18 patients. On admission, abnormalities in chest x-ray images were detected in 6 (33.3%) patients who had pneumonia. The mean duration from the first hospital admission to discharge was 33.1 ± 9.2 days.

**Discussion:** The majority of COVID-19 patients are male. COVID-19 comorbidities were found in several patients. The main clinical symptoms of COVID-19 in this study were cough, sore throat, and fever. The abnormal laboratory finding in COVID-19 patients is anemia, an increase in AST and ALT levels, and chest x-ray images of pneumonia. All patients are in mild condition. The average length of hospital admission patients to discharge is more than 30 days.

**Conclusion:** Although all patients are in mild condition, the inability of a local laboratory to check for positive confirmation of COVID-19 makes the admission period of the patient in the hospital very long. The availability of RT-PCR tests at Abdul Wahab Sjahranie Hospital Samarinda will greatly assist the further management of COVID-19 patients.

## Introduction

Coronavirus disease 2019 (COVID-19) is caused by severe acute respiratory syndrome coronavirus 2 (SARS-CoV-2) infection. SARS-CoV-2, an RNA virus from the Coronaviridae family, can be transmitted to humans and animals (1). Although most coronavirus infections in humans are mild, there have been two epidemics of beta coronavirus throughout history, namely severe acute respiratory syndrome (SARS) (2) and Middle East respiratory syndrome (MERS) (3), which have caused more than 10,000 cases in the last two decades, with a mortality rate of 10% for SARS (4) and 37% for MERS (5).

In December 2019, a series of pneumonia cases with unknown causes emerged in Wuhan, China, in which the clinical findings are similar to viral pneumonia. (6). The sequencing analysis from respiratory tract samples indicated a new coronavirus, which was then named SARS-CoV-2 (7). Until the end of May 2020, there are more than 5 million positive confirmed cases of COVID-19 worldwide, with the USA, Brazil, and Russia as countries for the most cases in the world (8).

Indonesia officially established its first COVID-19 confirmation case on March 2, 2020 from the capital of Jakarta (9). Meanwhile the first case of COVID-19 from East Kalimantan, a province of Indonesia in Borneo island, was reported on March 18, 2020, which was a patient from Abdul Wahab Sjahranie Hospital in Samarinda city (10).

East Kalimantan province has been determined as a candidate for the new capital of the Republic of Indonesia by 2019 (11). Therefore, the flow of people in and out of East Kalimantan has been increasing rapidly, especially in Samarinda as the capital of the province. This movement of people plays an important role in the spread of COVID-19 in Samarinda. Abdul Wahab Sjahranie Hospital, the largest hospital in Samarinda, has been designated as the main referral hospital for COVID-19 patients in Samarinda (12). Currently, there is no previous study has been done which describe the COVID-19 cases from East Kalimantan. Therefore, the study aims to describe the epidemiological, clinical, laboratory, and radiological characteristics of COVID-19 patients who are admitted at Abdul Wahab Sjahranie Hospital Samarinda.

## Methods

### Patients

The subjects in this study were all patients admitted at Abdul Wahab Sjahranie Hospital with laboratory confirmation of positive COVID-19 results from nasopharyngeal swabs from March to May 2020. This study was approved by the Ethical Health Research Commission of Abdul Wahab Sjahranie Hospital (No. 070/Diklit/1328/IV/2020).

### Procedures

Nasopharyngeal swab samples were taken when the patient was admitted to the hospital, then the samples were sent to the Center for Health Laboratory in Surabaya, Indonesia as an official examination laboratory for COVID-19 by the Ministry of Health of the Republic of Indonesia.

Nasopharyngeal swabs samples were then examined to confirm COVID-19 using RT-PCR according to guidelines from the Ministry of Health of the Republic of Indonesia. A positive COVID-19 patient with clinical improvement can be discharged from the hospital if the follow-up results of RT-PCR examination from nasopharyngeal swab samples of two days in a row show negative results.

### Data collection

This study collected data on the hospital medical records of all patients with positive confirmation of COVID-19. Data was taken from patients admitted to the hospital from March 14, 2020 to May 2, 2020. Epidemiological, clinical, laboratory and radiological data of patients were reviewed with standardized data collection forms from hospital medical records. Two researchers independently reviewed data collection to double-check the data obtained.

### Statistical analysis

Continuous variables were expressed as mean ± SD; categorical variables were expressed as frequency and percentage. Statistical analyses were done using Microsoft Excel.

## Results

By May 8, 2020, 18 admitted hospital patients had been identified as having laboratory-confirmed COVID-19. Five (27.8%) of the COVID-19 patients were aged 30-39 years, and 4 (22.2%) were aged 60-69 years. The mean age of the patients was 44 ± 15.5 years. Most of the infected patients were men (16 [88.9%]); less than half had underlying diseases (7 [38.9%]), including hypertension (4 [22.2%]), cardiovascular disease (2 [11.1%]), and diabetes (1 [5.56%]).

The most common symptoms at onset of illness were cough (16 [88.9%] of 18 patients), sore throat (8 [44.4%]), and fever (8 [44.4%]); less common symptoms were runny nose (7 [38.9%], myalgia or fatigue (6 [33.3%]), dyspnea (5 [27.8%]), headache (4 [22.2%]), and nausea (4 [22.2%]). The mean duration from first hospital admission to COVID-19 laboratory confirmation was 10 ± 3.6 days, and to hospital discharge was 33.1 ± 9.2 days.

The blood counts of patients on admission showed leukocytosis (white blood cell count more than 10.8×10^9^/L; 2[11.1%] of 18 patients), lymphopenia (lymphocyte count < 19%; 3 [16.7%] patients), neutrophilia (neutrophil count > 74%; 3 [16.7%] patients), and monocytosis (monocytes count > 9%; 3 [16.7%] patients). Laboratory findings of patients on admission also showed low hemoglobin level (hemoglobin < 14 g/L; 6 [33.3%] patients), low erythrocytes count (red blood cell count < 4.7 x10^6^ per L; 5 [27.8%] patients), and high platelet large cell ratio (P-LCR > 25 g/L; 4 [22.2%] patients). All patients had high platelet distribution width (PDW > 13 g/L) on admission. Levels of aspartate aminotransferase (AST) and alanine aminotransferase (ALT) were increased in 10 (55.6%) of 18 patients. All patients had normal serum levels of procalcitonin on admission (0.26 ± 0.1 u/L). On admission, abnormalities in radiographic images were detected in 6 (33.3%) patients which had pneumonia.

As of May 23, 2020, all patients have been discharged. Patients with clinical improvement can be discharged from the hospital if the results of RT-PCR examination two days in a row show negative results.

**Table 1.** Demographics and baseline characteristics of COVID-19 patients.

| Characteristics | Mean ± SD or N (%) |
| --- | --- |
| Age, years | 44 ± 15.5 |
| 20-29 years | 3 (16.7) |
| 30-39 years | 5 (27.8) |
| 40-49 years | 3 (16.7) |
| 50-59 years | 3 (16.7) |
| 60-69 years | 4 (22.2) |
| Sex |  |
| Men | 16 (88.9) |
| Women | 2 (11.1) |
| Sources of transmission |  |
| Imported cases from Gowa, South Sulawesi | 12 (66.7) |
| Imported cases from other places | 6 (33.3) |
| Local transmission within Samarinda | 0 (0) |
| History of comorbidity | 7 (38.9) |
| Hypertension | 4 (22.2) |
| Cardiovascular disease | 2 (11.1) |
| Diabetes mellitus | 1 (5.6) |
| BMI, kg/m<sup>2</sup> | 23.7 ± 2.8 |
| BMI > 25 kg/m<sup>2</sup> | 5 (27.8) |

**Table 2.**
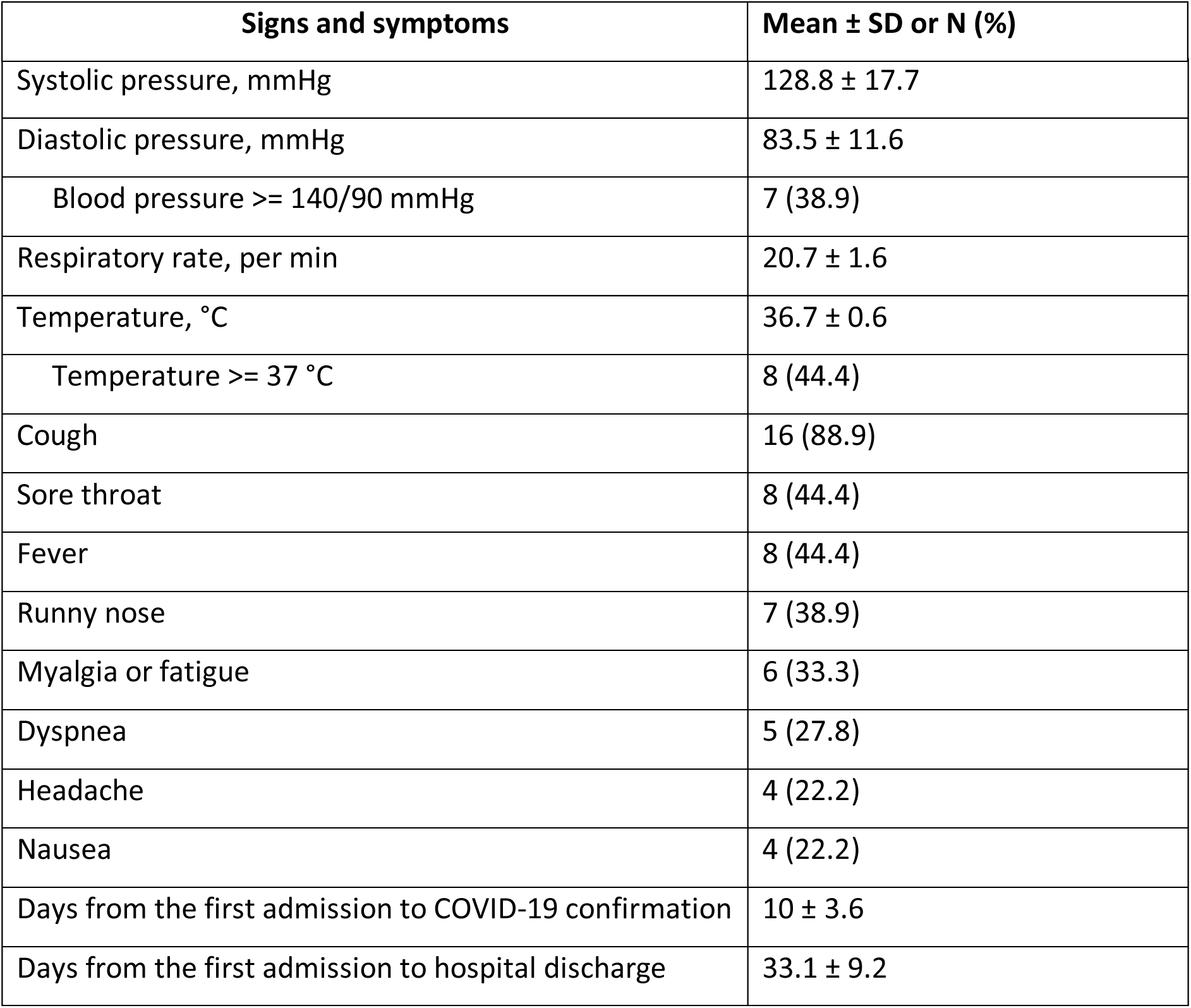
Signs and symptoms characteristics of COVID-19 patients.

| Signs and symptoms | Mean ± SD or N (%) |
| --- | --- |
| Systolic pressure, mmHg | 128.8 ± 17.7 |
| Diastolic pressure, mmHg | 83.5 ± 11.6 |
| Blood pressure ≥ 140/90 mmHg | 7 (38.9) |
| Respiratory rate, per min | 20.7 ± 1.6 |
| Temperature, °C | 36.7 ± 0.6 |
| Temperature ≥ 37 °C | 8 (44.4) |
| Cough | 16 (88.9) |
| Sore throat | 8 (44.4) |
| Fever | 8 (44.4) |
| Runny nose | 7 (38.9) |
| Myalgia or fatigue | 6 (33.3) |
| Dyspnea | 5 (27.8) |
| Headache | 4 (22.2) |
| Nausea | 4 (22.2) |
| Days from the first admission to COVID-19 confirmation | 10 ± 3.6 |
| Days from the first admission to hospital discharge | 33.1 ± 9.2 |

**Table 3.**
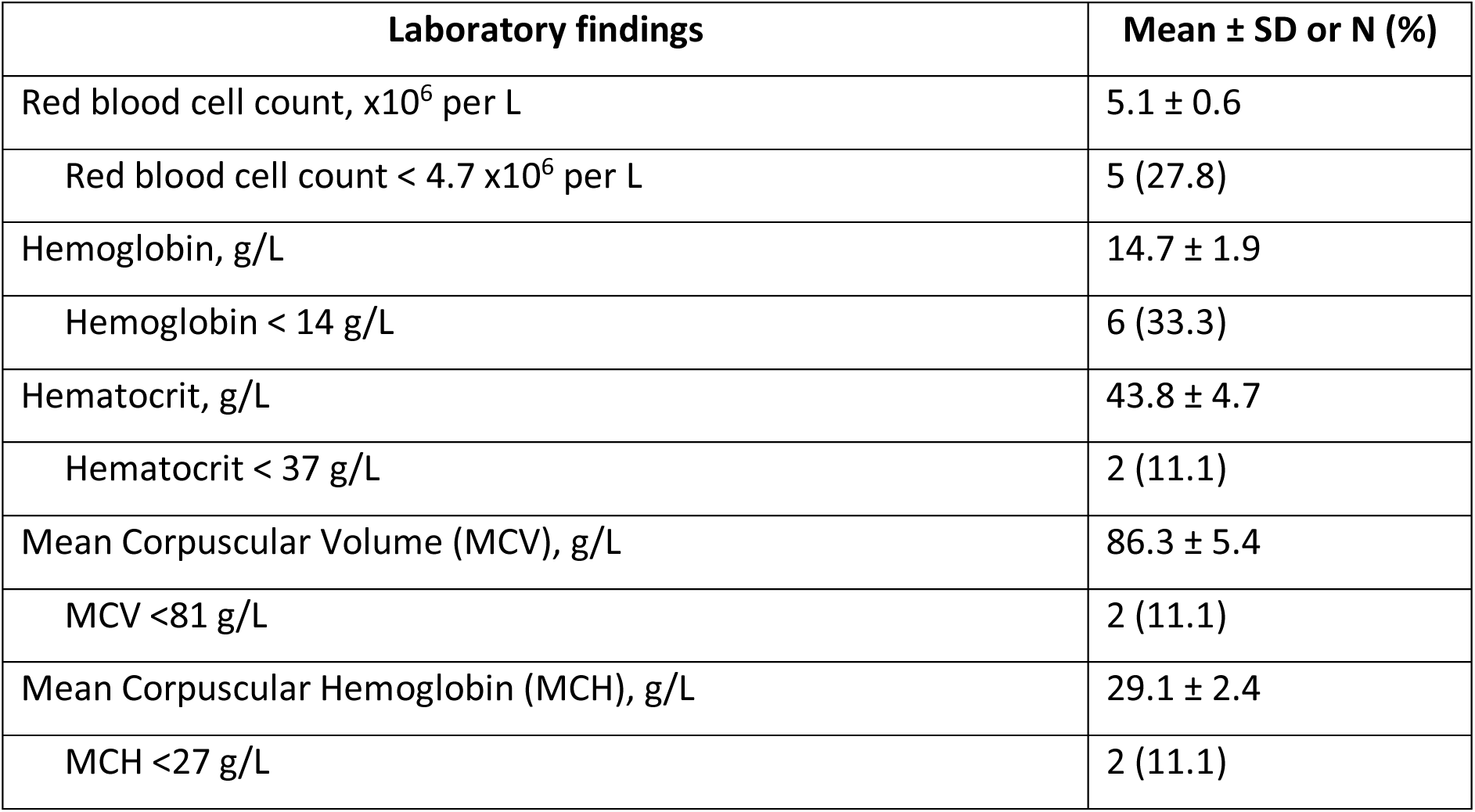
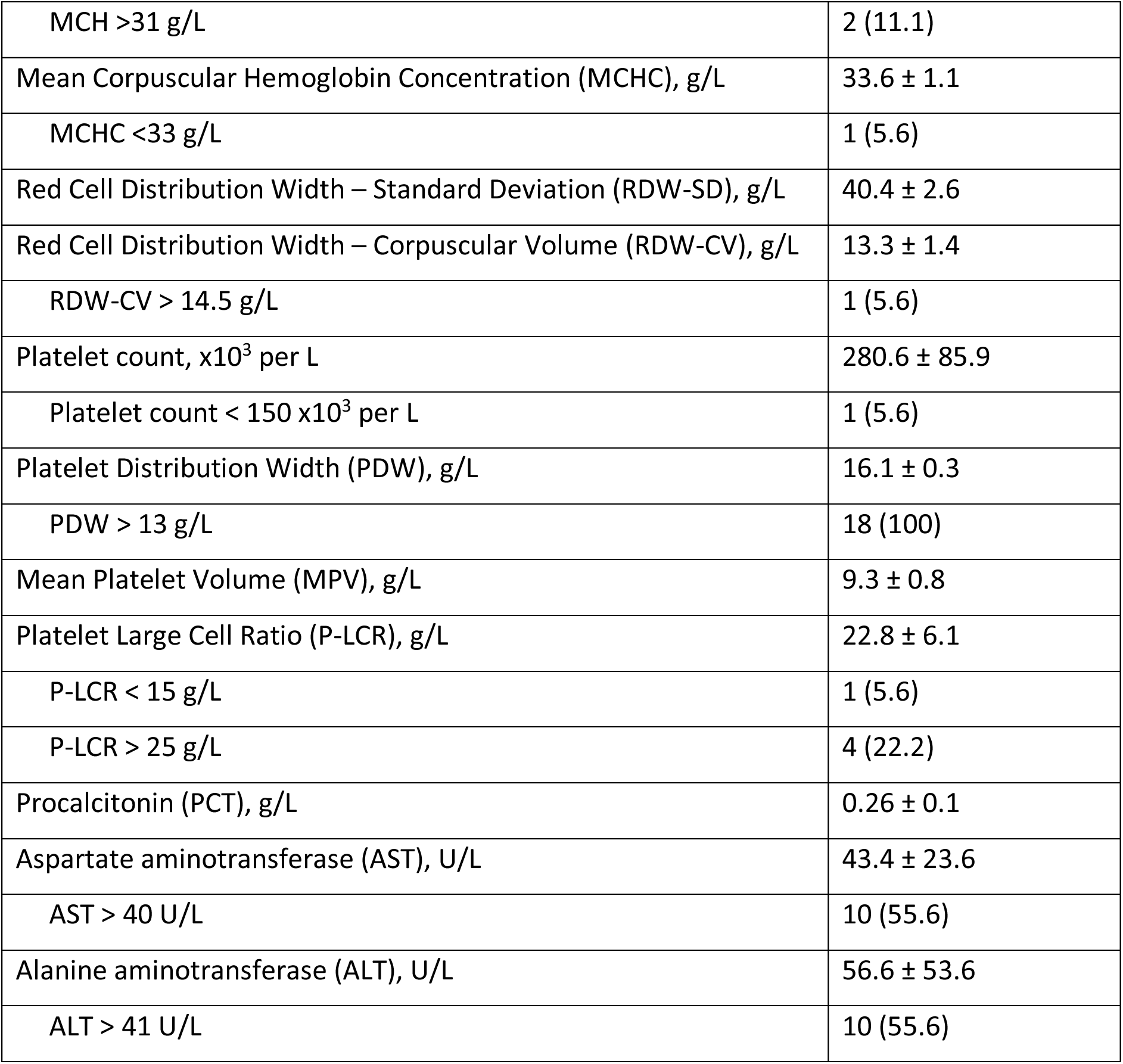
Laboratory findings of COVID-19 patients on hospital admission.

**Table 4.** White blood cell laboratory findings of COVID-19 patients on hospital admission.

| Laboratory findings | Mean ± SD or N (%) |
| --- | --- |
| White blood cell count, x10<sup>3</sup> per L | 7.8 ± 2.3 |
| White blood cell count > 10.8 x10<sup>3</sup> per L | 2 (11.1) |
| Neutrophil count, x10<sup>9</sup> per L | 4.8 ± 2.2 |
| Neutrophil count > 7 x10<sup>9</sup> per L | 2 (11.1) |
| Neutrophil count, % | 59.9 ± 13.4 |
| Neutrophil count > 74% | 3 (16.7) |
| Neutrophil count < 40% | 1 (5.6) |
| Lymphocytes count, x10<sup>9</sup> per L | 2.3 ± 1.1 |
| Lymphocytes count, % | 30.6 ± 12.3 |
| Lymphocytes count > 48% | 1 (5.6) |
| Lymphocytes count < 19% | 3 (16.7) |
| Monocytes count, x10<sup>9</sup> per L | 0.5 ± 0.1 |
| Monocytes count, % | 6.9 ± 1.9 |
| Monocytes count > 9% | 3 (16.7) |
| Eosinophils count, x10<sup>9</sup> per L | 0.2 ± 0.2 |
| Eosinophil count > 0,8 x10<sup>9</sup> per L | 1 (5.6) |
| Eosinophils count, % | 2.7 ± 2.4 |
| Eosinophil count >7% | 1 (5.6) |

**Table 5.**
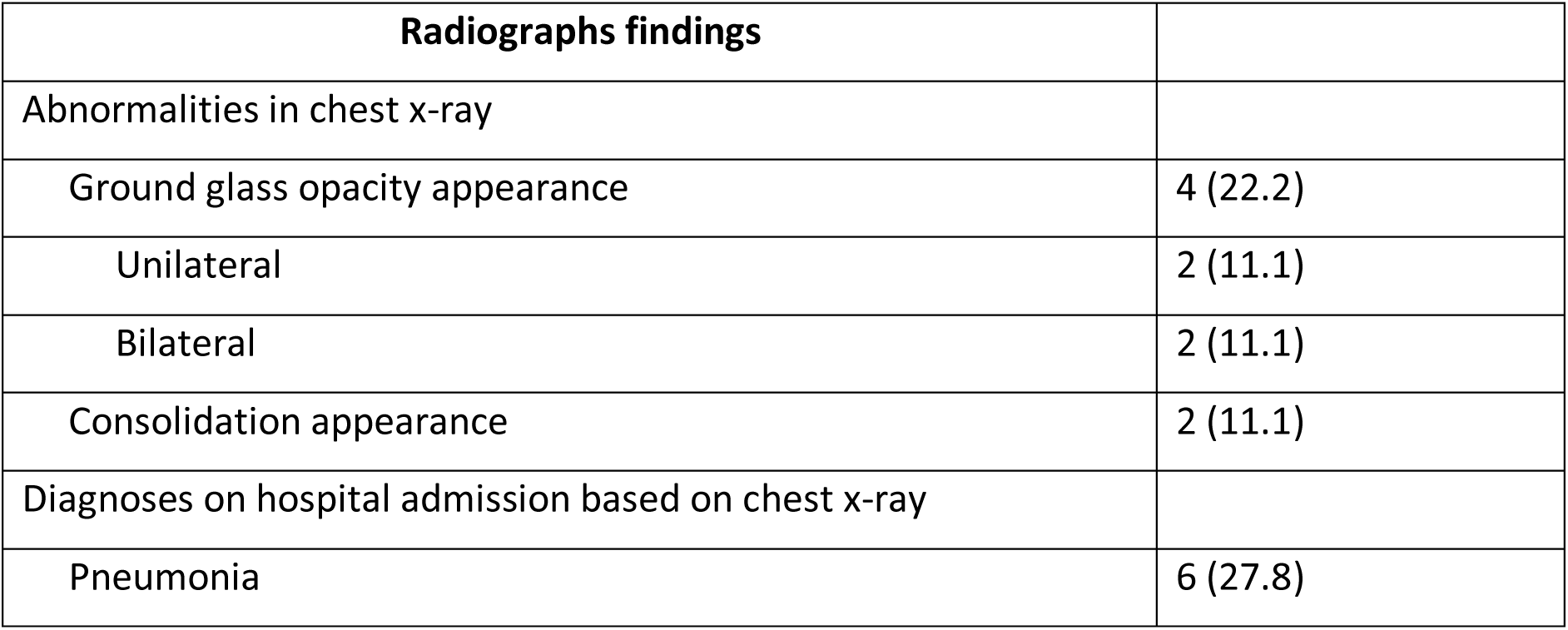
Radiographs findings of COVID-19 patients on hospital admission.

## Discussion

This study reports of 18 patients with positive COVID-19 confirmations admitted at Abdul Wahab Sjahranie Hospital Samarinda, Indonesia. As of May 24, 2020, there have been 276 positive COVID-19 positive confirmation cases in East Kalimantan (13). There are 22,271 positive confirmed cases of COVID-19, with 5,402 people recovering and 1,372 patients have died in Indonesia on May 24, 2020 (14).

The majority of COVID-19 patients in Abdul Wahab Sjahranie Hospital are male. This is consistent with a meta-analysis study which showed that men took the largest percentage in the distribution of COVID-19 according to gender (15). Other studies have shown that SARS-CoV and MERS-CoV also infect more men than women (16). The low vulnerability of women to viral infections is thought to be caused by the protective effect of the X chromosome (17) and the effects of sex hormones such as estrogen and progesterone, which play an important role in the immune system (18).

COVID-19 comorbidities were found in several patients in this study, such as hypertension, cardiovascular disease, and diabetes. This is consistent with a systematic review study and meta-analysis that shows that the major comorbidities are hypertension and diabetes, followed by cardiovascular disease. (19). This chronic disease is related to the pathogenesis of COVID-19. Chronic disease has a standard picture similar to infectious diseases, especially in the immune system (20). Metabolic disorders can reduce immune function by inhibiting the work of macrophages and lymphocyte function, thus making a person more susceptible to complications from a disease (21).

The main clinical symptoms of COVID-19 in this study were cough, sore throat, and fever. This is consistent with a meta-analysis study showing that the main clinical symptoms of COVID-19 patients are fever and cough (15). COVID-19 patients generally present with similar symptoms, such as fever, cough, and sore throat. Most patients with SARS-CoV-2 infection will appear with mild flu-like symptoms (22).

Several COVID-19 patients had anemia, which is a low level of hemoglobin and red blood cell count. This can be caused by the initial condition of the patient at the hospital who already has anemia. Iron deficiency is a form of anemia that is common in patients admitted to hospitals in Indonesia (23). A study found that iron deficiency anemia can affect the immune response and cytokine activity in the body (24). This can increase the vulnerability to be infected with COVID-19.

Another abnormal laboratory finding in COVID-19 patients here is an increase in AST and ALT levels which are parameters of liver function. This is similar to other studies that the laboratory examination results of COVID-19 patients showed that 76.3% had impaired liver function (25). Impaired liver function related to COVID-19 can be caused directly by a viral infection or other conditions such as the use of drugs that are hepatotoxic or due to systemic inflammatory responses, hypoxic conditions due to respiratory distress, and multiorgan dysfunction (26). If impaired liver function occurs, hepatoprotective drugs are recommended to be given to these patients (27).

Abnormal chest x-ray images of pneumonia were only found in 6 patients in this study. This is similar to other studies that found that consolidation and ground-glass opacity are chest x-ray findings in patients with positive confirmation of COVID-19 (28). The description of bilateral pneumonia with ground-glass opacity appearance in this study is consistent with the results of a systematic review and meta-analysis of COVID-19 (29).

All patients are in mild condition and none of the patients admitted to intensive care. This is consistent with studies that estimate the number of asymptomatic patients for COVID-19 around less than half (30). As of May 24, 2020, only 3 fatal cases of laboratory-confirmed COVID-19 were reported in East Kalimantan (13). Most of the COVID-19 patients admitted to hospitals in East Kalimantan are in mild condition.

The average length of time a patient starts in the hospital until the laboratory positive COVID-19 is confirmed is more than 10 days. This is due to the nasopharyngeal swab sample from Abdul Wahab Sjahranie Hospital being sent to the Center for Health Laboratory of the Ministry of Health in Surabaya, Indonesia. This laboratory serves the examination of reference specimens from the provinces of East, Central, South, and North Kalimantan (12). As of May 24, 2020, there were 276 positive cases in East Kalimantan, 308 in Central Kalimantan, 599 in South Kalimantan, and 164 North Kalimantan (31). The large number of samples that must be examined up to 4 provinces makes the slow progress of laboratory examination. Patients must wait a long time for laboratory confirmation results of COVID-19.

The average length of hospital admission patients to discharge is more than 30 days. This is because COVID-19 patients with clinical improvement can only be discharged from the hospital if the results of RT-PCR examination two days in a row show negative results (32). Based on the explanation above, the large number of samples that had to be examined from 4 provinces in Kalimantan by the Center for Health Laboratory of the Ministry of Health made patients wait longer for negative COVID-19 confirmation results.

Our study has several limitations. First, with a limited number of cases, this is a series of small patient cases. Data collection for larger studies will help to better determine the clinical picture and risk factors for this disease. Second, there is a potential exposure bias and no children were reported as patients in this study. More efforts must be made to answer these questions in further research.

## Conclusion

All COVID-19 patients in this study were in mild condition with symptoms of fever, cough, and sore throat. Most patients were male and had comorbidities which can increase the risk of COVID-19. Anemic conditions and increased AST/ALT levels in some patients need attention during patient care in the hospital. Although all patients are in mild condition, the inability of a local laboratory in East Kalimantan to check for positive confirmation of COVID-19 extended the admission period of the patient in the hospital unnecessarily. Therefore, the availability of RT-PCR tests at Abdul Wahab Sjahranie Hospital Samarinda will greatly assist the further management of COVID-19 patients.

## Data Availability

The data that support the findings of this study are available from the corresponding author, SP, upon reasonable request.

## Notes

### Competing Interest Statement

The authors have declared no competing interest.

### Funding Statement

Funding from the Ministry of Education and Culture. Faculty of Medicine Mulawarman University, Samarinda, Indonesia. The funder of the study had no role in study design, data collection, data analysis, data interpretation, or writing of the report.

### Author Declarations

This study was approved by the Ethical Health Research Commission of Abdul Wahab Sjahranie Hospital (No. 070/Diklit/1328/IV/2020).

